# Modelling of immune response in chronic myeloid leukemia patients suggests potential for treatment reduction prior to cessation

**DOI:** 10.1101/2022.06.28.22277004

**Authors:** Elena Karg, Christoph Baldow, Thomas Zerjatke, Richard E Clark, Ingo Roeder, Artur C. Fassoni, Ingmar Glauche

**Affiliations:** Institute for Medical Informatics and Biometry, Carl Gustav Carus Faculty of Medicine, Technische Universität Dresden, Dresden, Germany; Department of Molecular and Clinical Cancer Medicine, University of Liverpool, Liverpool, UK; National Center for Tumor Diseases (NCT), Dresden, Germany: German Cancer Research Center (DKFZ), Heidelberg, Germany; Faculty of Medicine and University Hospital Carl Gustav Carus, Technische Universität Dresden, Dresden, Germany; Helmholtz-Zentrum Dresden–Rossendorf (HZDR), Dresden, Germany; Instituto de Matemática e Computação, Universidade Federal de Itajubá, Itajubá, Brazil

## Abstract

Tyrosine kinase inhibitors (TKI) are highly efficient drugs to combat Chronic Myeloid Leukemia (CML). While long-term drug administration is associated with side effects and high economic costs, current efforts focus on the discontinuation of TKI therapy for well-responding patients. The DESTINY trial showed that TKI dose reduction prior to cessation can lead to an increased number of patients achieving sustained treatment free remission (TFR). However, there has been no systematic investigation to evaluate how dose reduction regimens can further improve the success of TKI stop trials.

Here, we apply an established mathematical model of CML therapy to investigate different TKI dose reduction schemes prior to therapy cessation and evaluate them with respect to the total amount of drug used and the expected TFR success. Our analysis is based on a cohort of 72 patients from the DESTINY trial for which we obtained individual model fits.

Our systematic analysis confirms clinical findings that the overall time of TKI treatment is a major determinant of TFR success, while highlighting that lower dose TKI treatment for the same duration is equally sufficient for many patients. Applying the model to an unstratified cohort of CML patients in deep remission our results suggest that a stepwise dose reduction prior to TKI cessation can increase the success rate of TFR, while substantially reducing the amount of administered TKI.

Our findings illustrate the potential of dose reduction schemes prior to treatment cessation and suggest corresponding and clinically testable strategies that are applicable to many CML patients.

**Summary:** Discontinuation of tyrosine kinase inhibitor (TKI) treatment is emerging as the main therapy goal for patients suffering from Chronic Myeloid Leukemia, although it is not clear how dose reduction regimens can improve the achievement of treatment free remission (TFR). Our systematic analysis uses mathematical models of leukemia-immune interaction to show that a stepwise dose reduction prior to TKI cessation can potentially increase the success rate of TFR, while substantially reducing the amount of total administered TKI.

## Introduction

Chronic myeloid leukemia (CML) is a malignancy of the hematopoietic stem cell. The introduction of tyrosine kinase inhibitors (TKI) revolutionized CML treatment as they target the causative BCR-ABL1 oncoprotein (1). Most patients respond well to TKI treatment and achieve sustained molecular remission, defined as low levels of *BCR-ABL1* mRNA (2, 3). Optimally responding patients treated with the first generation TKI imatinib achieve a 3-log reduction in *BCR-ABL1* levels (denoted as major molecular remission, MR3) after a median of 18 months and a 4-log reduction (MR4) after a median of 45 months (4). Related to this molecular response, the overall life expectancy of CML patients approaches that of a healthy reference cohort (5). However, continuous treatment with TKI can impose adverse effects and is costly (6). Therefore, several studies have investigated whether optimally responding patients can stop TKI therapy yet remain in treatment free remission (TFR) (7, 8). Consistently, about 50% of patients can achieve sustained TFR while 50% develop molecular disease recurrence, typically within two years of stopping (8-11). Although longer treatment duration and deep molecular remission prior to stopping are favorable prognostic markers of TFR (8), it is still not possible to prospectively identify patients likely to undergo disease recurrence and thus exclude them from TFR attempts.

The reasons why some patients remain in TFR while others present with molecular recurrence are not clear. Complete eradication of CML cells during TKI therapy is unlikely, as this occurs over much longer time scales (12, 13), if at all (14). Moreover, some patients in sustained TFR still have measurable residual *BCR-ABL1* levels, suggesting that other factors sustainably control the remaining leukemic cells (15). There is accumulating evidence that the immune system contributes to this control (16-20), and there is evidence that TKI-driven reduction of leukemic cells shifts this balance and reactivates immune surveillance in CML (21, 22). This effect is further modulated once TKI treatment stops and might well influence the success of TFR (16, 23).

The integration of treatment data with underlying mathematical models can address the structural conditions necessary for a stable balance between leukemia remission and immunological control. We and others have shown that a bidirectional interaction between leukemia growth and anti-leukemic effects, such as a specific immune response, is a prerequisite to obtain stable remission scenarios (24-26). It remains to be investigated to what extent an adapted TKI treatment before stopping can maintain molecular remission while sufficiently stimulating the immune system to establish long term immune surveillance.

In this context, the DESTINY trial is of particular interest, as it specifically alters the TKI treatment schedule before patients stop TKI (27). Its study protocol includes a 12 month TKI reduction period to 50% of the original dose prior to complete cessation, which improved TFR rates to > 60%. We previously demonstrated that the initial dose reduction period is indeed informative to identify a subset of patients with a high risk for TFR failure (28), by showing that 87.9% of patients with highly increasing *BCR-ABL1* values during this time experienced a molecular recurrence, compared with only 27.5% recurrence in the group with no or low increase of *BCR-ABL1*. From this we concluded that rapidly increasing *BCR-ABL1* values during dose reduction strongly suggest TFR failure after TKI stopping.

The results of the DESTINY trial also raise the question whether other dose reduction strategies prior to stopping could further increase the overall success rate of TFR. To address this, we have adapted our previously suggested mathematical model of CML treatment and immune response (29) to describe extended CML time course data from the DESTINY trial. Thereby we obtain an indirect estimate of the leukemia-immune interaction of a larger patient cohort, which we further use to illustrate a novel modeling strategy allowing us to explore whether amended schedules of TKI application, including different schedules of dose reduction, can influence the overall success rate of TFR.

## Methods

### Patient data

Our mathematical modelling approach is based on patient data from the DESTINY trial (NCT 01804985), whose primary results have been previously reported (27). The trial studied the effect of TKI reduction to 50% of the standard dose for 12 months prior to TKI cessation. All patients were previously treated with TKI monotherapy (either with imatinib, dasatinib or nilotinib) for at least three years and achieved stable molecular remission (MR3 in 49 patients and MR4 in 125 patients) for at least 12 months prior to entering the trial (see Supplementary Text S1, Supplementary Figure S1A). For the numerical analysis we use a logarithmic transformation of the *BCR-ABL1/ABL1* ratios (LRATIO = log10(*BCR-ABL1/ABL1*)).

In order to adapt the mathematical model to informative patient time courses, we applied several selection criteria to ensure that sufficient measurements were available during early TKI treatment, dose reduction and follow up (Figure 1A). This yields a total of 72 time courses for a detailed analysis (denoted as *clinical reference data set*). Initial statistical assessment revealed no differences between the original patient cohort and the reference data set with respect to initial LRATIO, treatment duration, follow-up duration, recurrence times and type of TKI (Supplementary Figure S1B-G). The selection process moderately increases the proportion of CML recurrences from 38.5% in the original to 51.4% in the selected cohort.

**Figure 1:**
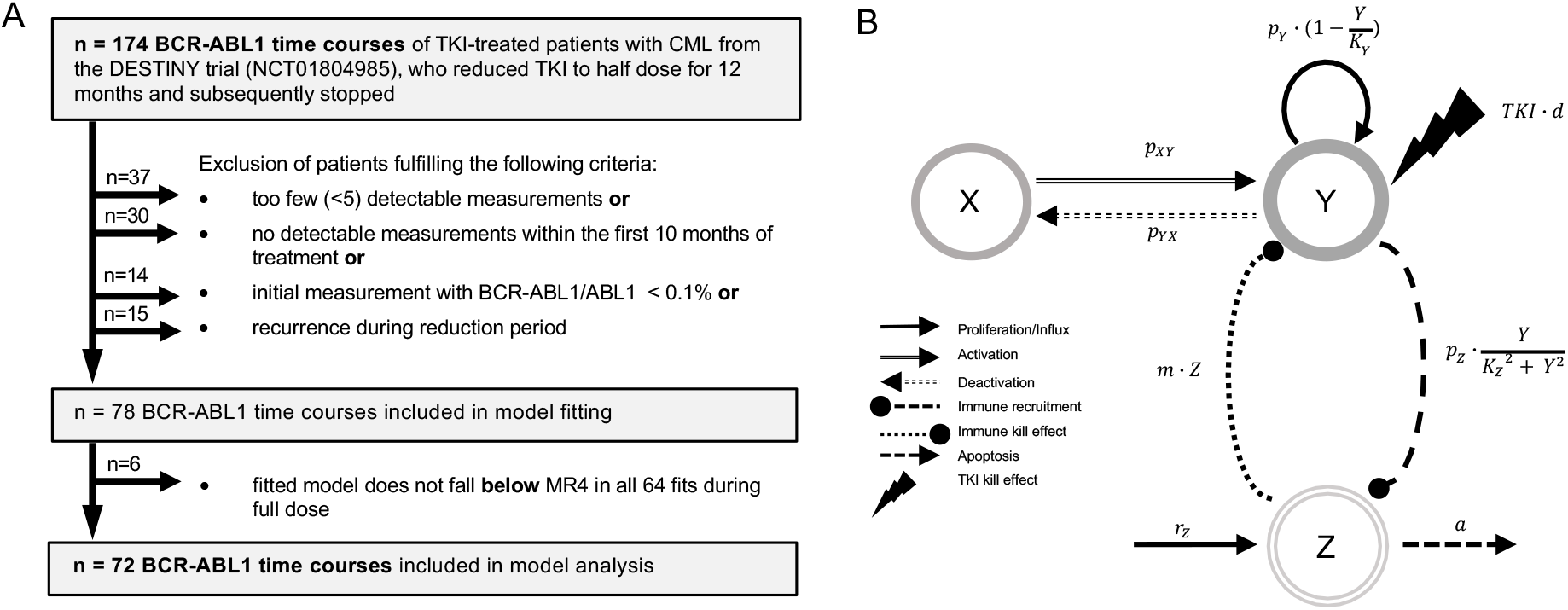
**A)** Flow diagram indicating the strategy for patient selection. Patients were excluded with less than 5 detectable BCR-ABL1 measurements, no detectable measurements, or an initial BCR-ABL1/ABL1-ratio below 0.1%. Furthermore, we excluded patients that presented with a recurrence during reduction period. After model fitting, we identified and excluded 6 patients, for which not all of the 64 simulation fits did predict BCR-ABL1 levels below MR4 during the full dose treatment period. **B)** General scheme of the ODE model setup indicating the relevant cell populations and their mutual interactions (arrows with rate constants) that govern their dynamical responses. Leukemic cells can reversibly switch between the quiescent (*X*) and proliferating (*Y*) state with corresponding transition rates *p*_*XY*_ and *p*_*YX*_. Proliferating cells divide with rate 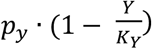. The TKI treatment has a cytotoxic effect *TKI* on proliferating cells (lightning symbol, linearly depending on the dose *d*) while quiescent cells are not affected. Immune cells in *Z* have a cytotoxic effect (with rate *m*) on proliferating leukemic cells in *Y*. The proliferation of immune cells is stimulated in the presence of proliferating leukemic cells by an immune recruitment rate 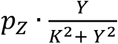. This nonlinear term describes an immune window, where the immune response is suppressed for high leukemic cell levels above the constant *K*_*Z*_. Moreover, immune cells are generated by a constant production *r*_*Z*_ and undergo apoptosis with rate *a* (see Materials and Methods).

### Mathematical model of TKI-treated CML

We describe individual disease dynamics of CML patients (i.e. time course of *BCR-ABL1/ABL1* ratios) in terms of a mathematical model which comprises interactions between immune effector cells (*Z*), quiescent (*X*) and proliferating (*Y*) leukemic stem cells (Figure 1B). Herein, we extend the previously introduced model (29) based on ordinary differential equations (ODEs), to account for the intermediate dose reduction period. Assuming a linear dose-response relationship for the TKI treatment (30), the dosage factor *d* declines from *d=1* (full dose) to *d=0*.*5* or *0*.*25* (reduced dose) to *d=0* (therapy cessation). Details of the model are provided in Supplementary Text S2.

For comparison of the model simulation results with clinical time courses of *BCR-ABL1/ABL1* ratios measured in peripheral blood, we calculate the simulated *BCR-ABL1/ABL1* ratio (in %) and its logarithm as

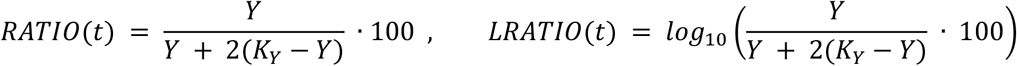

thereby accounting for the presence of both *BCR-ABL1* and *ABL1* transcripts in leukemic cells. Herein the carrying capacity *K*_*Y*_ is set to 10^6^. Within the model, we define molecular recurrence as the first time point at which RATIO increases above 0.1% and remains there for at least one month.

We obtain patient-specific optimal parameter choices for the model parameters (termed *λ*) by applying an optimization routine that minimizes the distance between each clinical time course and the corresponding simulation (see Supplementary Text S3).

## Results

### Model description of *BCR-ABL1* dynamics during dose reduction and after TKI cessation

We analyzed a cohort of 72 patients from the DESTINY trial (NCT01804985, (27)), for which complete time course information, i.e. *BCR-ABL1* measurements during the initial treatment response, during the 12-month dose reduction period, and after TKI stop, were available (*clinical reference data set*, Figure 1A). We hypothesized that these complete time courses reveal patient-specific features with respect to CML progression, TKI response and immunological control mechanisms. In order to quantitatively address these aspects, we applied an established mathematical model of CML treatment (12, 13, 31), which explicitly considers interactions between leukemic cells and the immune system (Figure 1B, see Methods) (29).

Figure 2 illustrates the general approach to obtain individual choices of model parameters to fit a patient’s time course. We applied a genetic algorithm to optimize the following seven critical parameters: the transition rates between quiescent and active LSC, *p*_*XY*_ and *p*_*YX*_, the proliferation rate of active LSC *p*_*Y*_, the TKI-dependent kill effect *TKI*, immune parameters *p*_*Z*_ and *K*_*Z*_, and the initial *BCR-ABL1/ABL1* ratio on the log-scale (denoted as *initLRATIO)*. While we cannot be sure whether a unique global optimum is identifiable, we observed that the measurement uncertainties allowed for many reasonable fits. For these reasons we took a complementary approach in which we explicitly consider the intrinsic heterogeneity of the model solutions. Technically, we used 64 independent optimization runs (*i* ∈ {1 … 64}) per patient *j*, and obtained 64 parameter sets *λ*_*ij*_ that reflect the patient-specific variability within the parameter space (Figure 2A, Supplementary Text S3). Figure 2B indicates the parameter choices for the particular example, while the final column depicts the residual error for each *λ*_*ij*_. Within the figure there are separate regions for some parameter choices (like the immune parameters *p*_*Z*_ and *K*_*Z*_), although the residual error remains in the same order of magnitude. This confirms the visual impression from Figure 2A that all 64 fits sufficiently explain the given patient time course (see Supplementary Figure S2 for further examples). Application of this optimization approach reassures us that all generic features of CML specific time courses, such as the initial biphasic response, sustained remission or recurrence can be reflected by our mathematical model.

**Figure 2:**
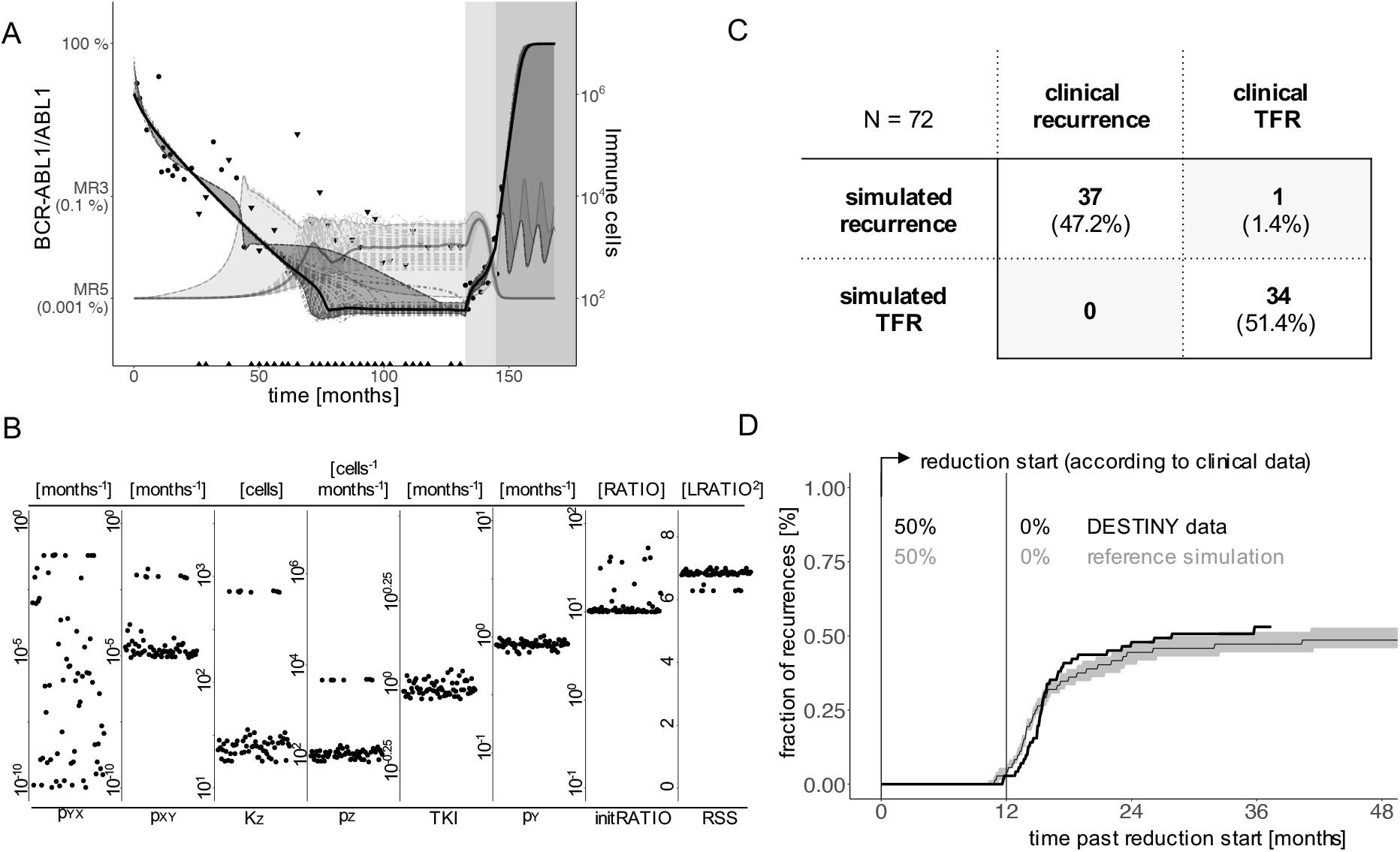
**A)** Example of clinical response data and model simulations of a patient with CML recurrence after therapy stop. Black dots indicate BCR-ABL1/ABL1-measurements; black triangles represent the quantification limit for undetectable BCR-ABL1 levels (see Supplementary Text S1). White background indicates full dose TKI treatment, light grey indicates the reduction period (50% of full dose), and dark grey refers to the time period after treatment cessation. The bold black line indicates the median and the dark grey ribbon corresponds to the 95% interval of all 64 simulated BCR-ABL1/ABL1-time courses (dashed dark grey lines). The bold grey line indicates the corresponding median of all 64 simulated immune cell counts (dashed light grey lines) with the light grey region corresponding to the 95% interval **B)** Distribution of individual parameter values of the 64 parameter sets of the same patient as in A, shown separately for the 7 parameters along with the residual sum of squares (RSS) as a measure of fitting quality. **C)** Comparison of the simulated results and the clinical outcomes with respect to TFR or disease recurrence according to a binary classification. Herein we define *simulated recurrence patient* if at least one of the 64 patient-specific parameter sets *λ*_*ij*_ resulted in a simulation with molecular recurrence during the follow-up period, while this is not the case for a patient classified as *simulated TFR*. **D)** Comparison of the time and fraction of recurrences between the patient data according to DESTINY (black line) and the reference simulation (grey). To obtain the curve for the reference simulation, we randomly choose one eligible parameter set *λ*_*ij*_ per each patient *j* and calculate whether and when there will be a recurrence in the corresponding simulation. We repeat this sampling approach 1000 times to obtain the median (grey line) and a 95%-confidence region (grey shaded). Time = 0 indicates the start of TKI dose reduction, which corresponds to the time point provided in the clinical data for each patient considered.

### Comparison of recurrence between model simulation and clinical data

Molecular recurrence within the DESTINY trial is defined as two consecutive *BCR-ABL1/ABL1* measurements which exceed 0.1% (MR3). In the model, we mirror this as sustained levels of leukemic cells above 0.1% for at least one month. Applying this definition to the simulations for each of the 64 parameter sets *λ*_*ij*_ for all 72 patients, we observed 34 patients with no indication of recurrence, while 38 have at least one simulation (i.e. one of the 64 parameter sets) predicting recurrence (including 10 with mixed outcomes and 28 with recurrence predictions only). Viewing this question as a binary classification problem, we applied a receiver operating characteristic (32) analysis to compare the simulated recurrences with true remission status from the *clinical reference data set* (Supplementary Figure S3). We obtained the best correspondence when the patients were classified as *simulated recurrence patient* if at least one of the 64 patient-specific parameter sets *λ*_*ij*_ resulted in a simulation with molecular recurrence during the follow-up period. Patients with only non-recurrence simulations were classified as *simulated TFR patients*. With this binary classification the model correctly reproduces the clinical outcome for nearly all patients (Figure 2C). While all 37 clinical recurrences are identified correctly by the model (37 true positives), one out of 35 cases with enduring TFR is missed by the model simulations.

In a complementary approach, we also include the timing of molecular recurrence events to compare our simulations with the *clinical reference data set* from the DESTINY trial. In order to account for the intrinsic heterogeneity of the model solutions we use a sampling approach in which for all 72 patients one of their eligible parameter set *λ*_*ij*_ is randomly chosen, while this whole process is repeated 1000 times (see Supplementary Text S4). We obtain the cumulative, time dependent incidence of recurrences as the median and the 95% range over the simulation results from all the repeated realizations. Figure 2D indicates that the model simulations well reflect the timing of molecular recurrences with a prominent increase within a few months after stopping TKI. The fraction of recurrences is slightly underestimated compared to the binary classification in Figure 2C as the repeated sampling strategy rather provides an average recurrence behavior per patient. We point out that for these reference simulations we are using the same time from TKI start until dose reduction as it is denoted for each individual patient in the DESTINY trial (on average 85.4 months).

Our results show that the suggested model is capable of reproducing *BCR-ABL1/ABL1* time courses of CML patients as well as the timing and occurrence of clinically observed recurrences. However, applying the same approach to pre-cessation data only, we show that the model cannot reliably predict the future remission status of a patient (Supplementary Figure S4, Supplementary Text S5). This is particularly true for patients with non- or slowly-increasing *BCR-ABL1* levels during dose reduction as an inference of functional immunological control cannot be obtained with sufficient precision. These findings complement a previous statistical analysis of the DESTINY data that obtained similar results (28).

### Immune landscapes account for within-patient heterogeneity

We have previously shown that the mathematical model implies different ‘immunological landscapes’ characterized by the existence or absence of typical steady states that can be obtained after therapy stop (25, 29). The occurrence, the number, and the status of these steady states are determined by the chosen model parameters *λ* and are, therefore, specific for each parameter fit (Figure 3A, see Supplementary Text S6). In brief, parameter sets belonging to *class A* are characterized by an insufficient immune response such that there is only one stable steady state describing leukemic dominance (E_High_). *Class B* refers to scenarios with a strong immune system in which the steady state for leukemic dominance (E_High_, similar to class A) is accompanied by a second steady state describing immune control of a sufficiently few leukemic cells (remission steady state, E_Low_). *Class C* presents with a similar immunological landscape as class B while the size of the remission steady state (E_Low_) is smaller and more difficult to achieve.

**Figure 3:**
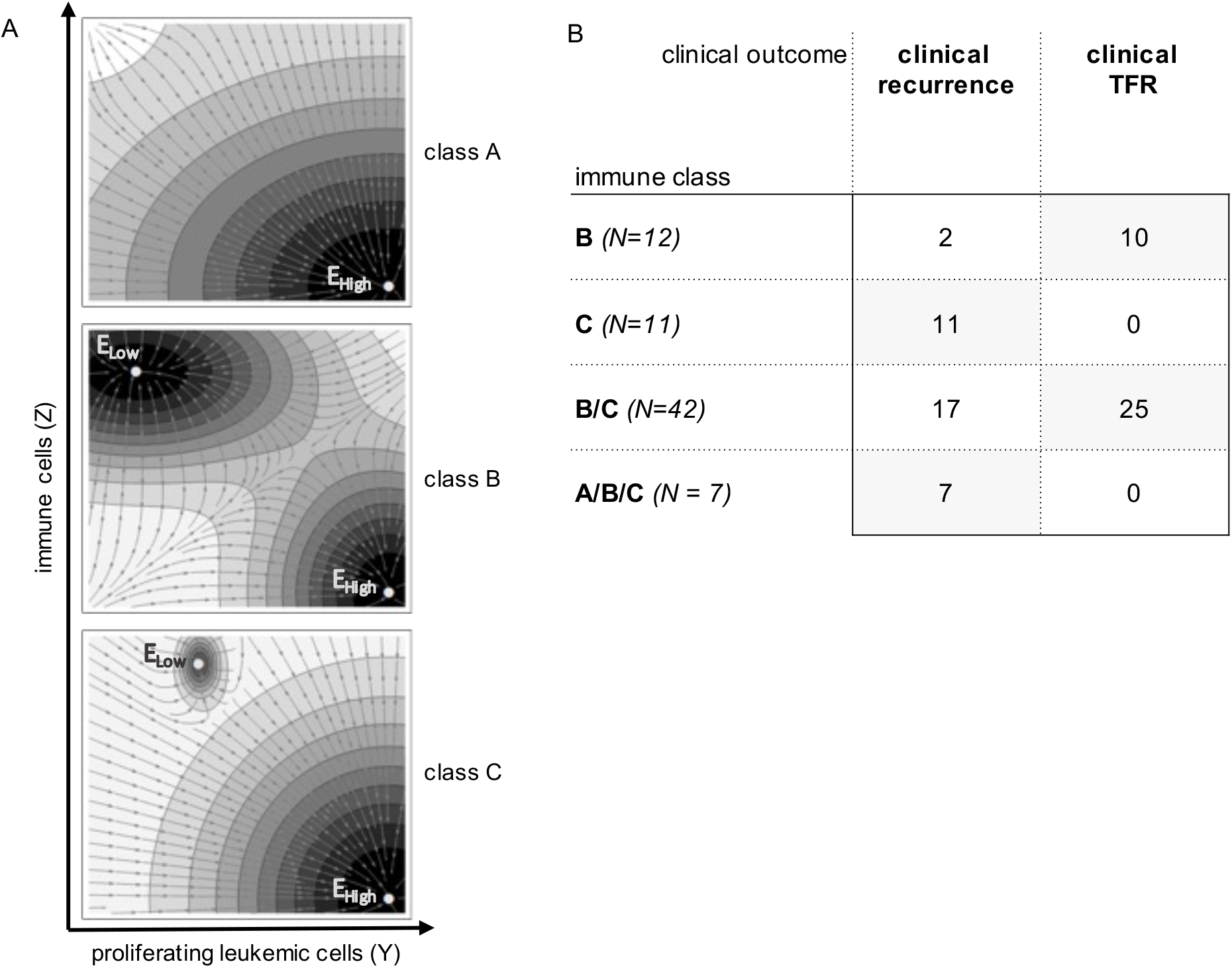
**A)** Sketch of the prototypical immunological landscape classes A, B and C (see Supplementary Text S6). The phase space is spanned by the number of proliferating leukemic cells *Y* on the x-axis and the number of immune cells *Z* on the y-axis. Steady states of the dynamical system are identified by E_High_ and E_Low_ (see Supplementary Text S6). The vector field indicates the direction of model trajectories after TKI cessation and the basin of attractions of the stable steady states. **B)** Classification of the 72 patients according to immune classes and their clinical outcomes after treatment cessation.

As we have not obtained a uniquely identifiable parameter set for all patients *j*, but use a spectrum of 64 optimization runs *i*, optimal parameter sets *λ*_*ij*_ of the same patient may be classified to different immunological classes A, B or C. Practically, each of the 64 optimal parameter configurations *λ*_*ij*_ for a particular patient *j*, obtained from fitting the complete data set (compare Figure 2A,B), corresponds to one of the three general immune classes A, B or C (Figure 3A). We observed 12 patients for which all the 64 fits were consistently identified as class B and 11 patients for which all fits were consistently identified as class C (Figure 3B, Supplementary Figure S2). However, there are 42 patients for which some parameter configurations indicated class B while others indicated class C (termed B/C). The same is true for 7 patients in which all three classes A, B and C were identified (termed A/B/C). Interestingly, class B patients were predominantly those achieving sustained TFR, while all of the class C patients developed molecular recurrence. The same is true for A/B/C patients, who all fail to achieve sustained TFR. B/C patients appear in both the molecular recurrence and in the sustained TFR groups. We will further analyze to which extend this intrinsic heterogeneity can explain how changes in the dose reduction schedule influence TFR success on the population level.

### Treatment duration determines overall TFR success

As a reliable prediction of the remission behavior for individual patients is limited by the insufficiency to infer immunological control parameters prior to TKI stopping, we aimed to investigate how general treatment schemes can be optimized such that they are applicable to all patients without prior stratification while maximizing TFR success and reducing TKI usage. We approach this question by comparing the temporal recurrence behavior (compare Figure 2D) and the average total TKI consumption for a range of systematically modified treatment schedules.

As a first step we simulated the 12 month dose reduction period starting strictly 24 months past reaching the 0.01% (MR4) remission level (MR4+24M+12M50% scenario) for all 72 patients instead of using the observed time points from the *clinical reference data set* (Figure 4A). Overall, the simulated CML recurrence for the amended treatment scheme is similar to that observed for the clinically reference simulation (Figure 4B). However, for the amended scenario, TKI is only administered for 24 months past reaching MR4, while the drug is given on average for 43.6 months after MR4 for the reference simulations (Figure 4A).

**Figure 4:**
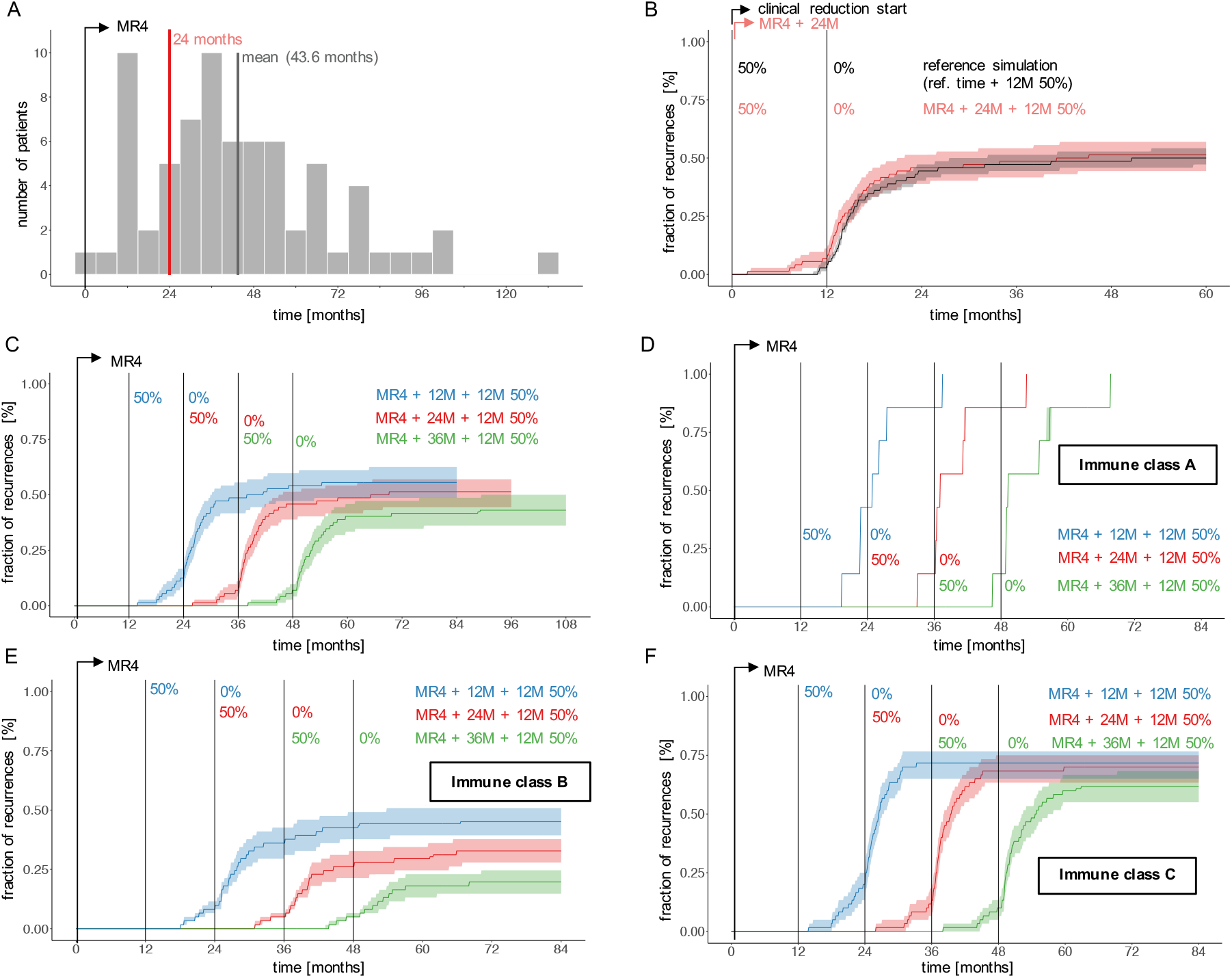
**A)** Histogram indicating the average time span per patient between reaching MR4 (time = 0) and start of TKI dose reduction for the clinical reference data from the DESTINY trial (grey bars) compared to the MR4+24M+12M50% scenarios in which dose reduction is initiated precisely 24 months after the corresponding fits reached MR4 (red line). **B)** Fraction of recurrences as a function of time comparing the reference simulation (grey; simulations according to the clinical reference data with respect to time point of dose reduction) and a scenario with a 12 month dose reduction period initiated 24 months after the simulated time courses have reached MR4 (red, indicated as MR4+24M+12M50%). Time = 0 corresponds to the start of TKI dose reduction, either according to the clinical data or of 24 months after reaching MR4. **C)** Same analysis in which the time between reaching MR4 and the initiation of the 12 month dose reduction period varies from 12 to 36 months (indicated as MR4+12M+12M50%, MR4+24M+12M50%, MR4+36M+12M50%). Time = 0 corresponds to the time point of reaching MR4 for each simulation. **D-F)** Corresponding simulations to subfigure C, stratified according to the immune class for each optimal parameter set *λ*_*ij*_. The subfigures only contain patients *j* with at least one optimal parameter set *λ*_*ij*_ classified to either immune class A, B, or C.

Varying the full dose TKI treatment time between 12 and 36 months past reaching MR4 (Figure 4C) clearly demonstrates that shorter TKI duration leads to more recurrences, while longer TKI treatment improves TFR success. The quantitative increase in TFR success per additional year of TKI treatment compares well with findings from the EURO-SKI trial on TKI discontinuation (8). To further examine how treatment duration acts on the different immune classes, we stratified the patients as to whether any of their optimal parameter sets *λ*_*ij*_ belong to class A, B or C. Applying the same sampling approach as above (Supplementary Text S4) separately to the eligible parameter sets of those subcohorts (Figure 4 D-F) indicated that both class B and class C configurations will benefit from longer treatments, although the benefit for class C is smaller. As expected, parameter configurations of class A develop molecular recurrence irrespective of the treatment configuration. Only patients that can in principle establish immune surveillance of their residual leukemia levels may therefore benefit from longer treatment durations.

### TKI dose reduction strategies to optimize TFR success

To address the effect of TKI dose on TFR success we considered a situation where patients receive 36 months full dose treatment past reaching MR4 (MR4 + 36M) and compared it to the reduction scheme discussed above, in which a 24 month full dose treatment after reaching MR4 precedes a 12 month reduction to either 50% of the original dose (MR4 + 24M + 12M50%) or to 25% of the original dose (MR4 + 24M + 12M25%). While some patients in the latter scenarios present with disease recurrence earlier than they would do with the full dose treatment, those patients are likely to fail anyway. Figure 5A indicates that those three scenarios can hardly be distinguished with respect to their long-term outcome, thereby suggesting that overall treatment duration determines the success rate, while there is potential to achieve the same results with substantially less TKI.

**Figure 5:**
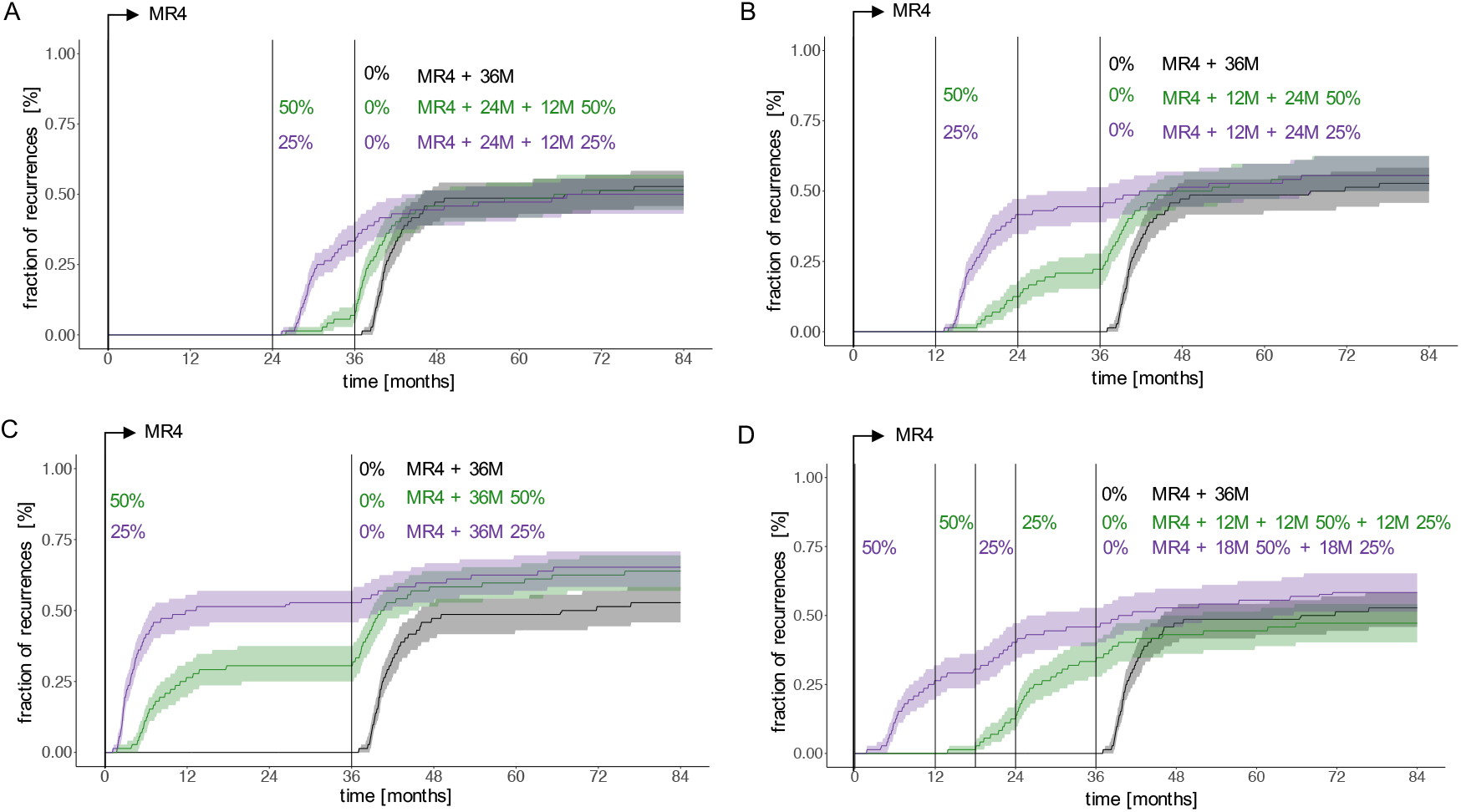
Fraction of recurrences as a function of time comparing immediate therapy cessation 36 months after reaching MR4 (black) with different scenarios of dose reduction covering the same overall treatment duration: **A)** 24 months full dose treatment past reaching MR4 plus 12 month dose reduction to 50% of the initial dose (green, indicated as MR4+24M+12M50%) and 24 months full dose treatment past reaching MR4 plus 12 month dose reduction to 25% of the initial dose (purple, indicated as MR4+24M+12M25%). **B)** 12 months full dose treatment past reaching MR4 plus 24 month dose reduction to 50% of the initial dose (green, indicated as MR4+12M+24M50%) and 12 months full dose treatment past reaching MR4 plus 24 month dose reduction to 25% of the initial dose (purple, indicated as MR4+12M+24M25%). **C)** dose reduction to 50% of the initial dose for 36 months immediately after reaching MR4 (green, indicated as MR4+36M50%) and dose reduction to 25% of the initial dose for 36 months immediately after reaching MR4 (purple, indicated as MR4+36M25%). **D)** 12 months full dose treatment past reaching MR4 plus 12 month dose reduction to 50% of the initial dose plus 12 month dose reduction to 25% of the initial dose (green, indicated as MR4+12M+12M50%+12M 25%) and dose reduction to 50% of the initial dose for 18 months immediately after reaching MR4 plus 18 month dose reduction to 25% of the initial dose (purple, indicated as MR4+18M50%+18M25%).

To systematically analyze this, we simulated further reduction schemes, all extending over 36 months past reaching MR4. Earlier initiation of dose reduction at 12 months past reaching MR4 leads to a similar fraction of recurrences compared to the above scenarios (MR4+12M+24M50% and MR4+12M+24M25%, Figure 5B). However, this effect is lost if the reduction step is initiated right after reaching MR4 (MR4+36M50% and MR4+36M25%, Figure 5C) resulting in an increase of the overall proportion of recurrences.

Complementary to this observation it is interesting that a stepwise dose reduction to 50% initially and 25% thereafter (MR4+12M+12M50%+12M25%) gives a lower recurrence rate (Figure 5D), while requiring only 58% of the TKI amount administered past MR4 compared to the full dose scenario. Again, an earlier introduction of the reduction regimen right after reaching MR4 (MR4+18M50%+18M25%) seems to oppose this effect. Having a closer look at the MR4+12M+12M50%+12M25% scenario we speculate that the advantage results from a better convergence of the immune class C patients to the remission steady state while this aim is not yet reached for patients of the immune class B (Supplementary Figure S5). For class C patients, full dose treatment may actually result in overtreatment and a deactivation of the immune system, while this effect is prevented by the stepwise reduction of TKI dose (Supplementary Figure S6). Our overall conclusions do not change if e.g. an overall treatment duration of 48 months post reaching MR4 is considered (Supplementary Figures S7).

## Discussion

Our results confirm clinical findings that the overall time of TKI treatment is a major determinant of TFR success (8), but at the same time indicate that lower dose TKI treatment may be sufficient to achieve the same results for many patients. Having a more detailed look on the response dynamics during dose reduction and after treatment cessation, we observe that a subset of patients with presumably insufficient immune control will inevitably relapse, no matter whether TKI is stopped all at once or reduced in a stepwise manner. During dose reduction, such patients often present with substantially increasing *BCR-ABL1* levels. However, in most patients there is no or only a mild increase of *BCR-ABL1* levels during TKI dose reduction, for which reliable predictions of TFR success after stopping treatment cannot be drawn (see also (28)). Our model results indicate that a stepwise dose reduction prior to TKI cessation does not limit the overall success rate of TFR for this patient group, while it can already substantially reduce TKI associated side effects (27, 33) as well as overall treatment costs.

The importance of the immune system in the sustained control of residual disease levels is widely recognized and many immunologically relevant subpopulations and their interactions are studied in the context of CML recurrence (21-23). While promising correlations, e.g. with the abundance of CD56^dim^ natural killer cells (18) or activated CD86^+^ plasmacytoid dendritic cells (19) have been identified, no unique marker nor control mechanism has been identified that allows a reliable prediction of TFR success. This is also supported by our dynamical analysis of CML time course data, which allows the identification of patients at high risk of molecular recurrence presenting with substantially increasing *BCR-ABL1* levels after TKI dose reduction but performs insufficiently to discriminate patients presenting with no or a mild increase of *BCR-ABL1* levels. However, our simulation approach allows us to study how an artificial, but individually parameterized patient cohort behaves for modified dose reduction schemes that can be applied to an unstratified patient cohort.

We explored the potential of TKI dose reduction beyond that used in the DESTINY trial. While we demonstrated that the initiation of 50% dose reduction 24 months after reaching MR4 closely mimics the heterogeneous timing of dose reductions from the DESTINY patients, we also explored more sophisticated dose reduction approaches. In particular, we observed that dose reductions to 25% of the original TKI dose also lead to comparable results. This finding also opens the possibility to initiate two-step reduction schemes that can further broaden the range of possible treatment options. Our simulations suggest that halving the dose twice in annual intervals one year after reaching MR4 (MR4 + 12M + 12M50% + 12M25%) could perform at least equally well compared to a full dose treatment with TKI over the whole three year period. We are looking forward to a recently initiated clinical trial investigating this question(32).

A range of clinical studies that administer TKI at lower than the standard dose underline the potential of these suggestions. Several studies, especially on second generation TKI in newly diagnosed patients, document the clinical efficacy of such reduced dose regimens to achieve cytogenetic and molecular remission while at the same time delivering an improved side effect profile (34-37). Other approaches use dose reduction for patients in major or deep molecular remission to manage TKI-related side effects and to improve the patients’ quality of life (38, 39). The results indicate that TKI dose reduction is safe, has no effect on long term outcome and only minimal effects on cytogenetic and molecular response (40). These clinical results are complemented by earlier mathematical modelling approaches of our group (31). In this work we could show that the cytotoxic effect of TKIs is limited by the rare activation of leukemic cells, thereby reasoning that higher than necessary TKI doses do not confer an additional benefit.

Our analysis along with the overall results of the DESTINY trial (27, 33) suggest that TKI dose reductions prior to stopping can confer an advantage with respect to TFR success compared to immediate TKI cessation. We hypothesize that this beneficial effect results from sensitization of the immune system prior to stopping. While it is beyond the scope of this work, these results stimulate speculations on the extent to which a mild increase in leukemic cell load under reduced TKI dose could be a useful trigger for immunological control mechanisms prior to stopping TKI. Further approaches to address this question may include vaccination strategies applied during deep remission (41) and before the patients discontinue therapy.

While both the small number of clinically available reference data sets and the simplifying assumptions underlying our modeling approach limit the generalizability of our results, we aimed for a strong internal consistency. To this end we used internal controls, such as the variation of overall TKI treatment durations, and compared them to clinical findings. Our results are, therefore, generic in nature but demonstrate a strong side of this systems biological approach: to underpin conceptual consideration with quantitative arguments that can guide the planning of experimental and clinical studies. For the particular situation, we argue that the clinical potential for TKI dose reductions in CML patients with sustained remission is not exhausted and does not compromise the goal of complete TKI cessation.

## Data Availability

The data used in this study are available upon request from the corresponding author.

## Ethical approval

The DESTINY trial ((27), NCT 01804985) was conducted in accordance with the Declaration of Helsinki and applicable regulatory requirements. The protocol was approved by the North West - Liverpool East Committee of the UK National Research Ethics Service.

## Data availability

The data used in this study are available upon request from the corresponding author.

## Authors’ Contributions

*Conception and design:* E. Karg, C. Baldow, I. Roeder, A.C. Fassoni, I. Glauche

*Development of methodology:* E. Karg, C. Baldow, T. Zerjatke. A.C. Fassoni, I. Glauche

*Acquisition of data (provided animals, acquired and managed patients, provided facilities, etc*.): R.E. Clark

*Analysis and interpretation of data (e*.*g*., *statistical analysis, biostatistics, computational analysis):* E. Karg, C. Baldow, A.C. Fassoni, I. Glauche

*Writing, review, and/or revision of the manuscript:* E. Karg, C. Baldow, T. Zerjatke, R.E. Clark I. Roeder, A.C. Fassoni, I. Glauche

*Study supervision:* I. Roeder, A.C. Fassoni, I. Glauche

## Acknowledgments

This work was supported by the German Federal Ministry of Research and Education (BMBF, grant number 031A315 “*MessAge*”) to I.G. and ERA-Net ERACo-SysMed JTC-2 project “*prediCt*” (Project No. 031L0136A) to I.R. E.K. was funded via a José Carreras-DGHO-Promotionsstipendium and the Graduate academy of TU Dresden.

