## Supplementary Text and Figures for "Modelling of immune response in chronic myeloid leukemia patients suggests potential for treatment reduction prior to cessation"

#### S1: Patient cohort and *BCR-ABL1* measurements in the DESTINY trial

Our analysis is based on the DESTINY trial (NCT 01804985) [1]. All patients were previously treated with TKI monotherapy prior to entering the trial. *BCR-ABL1/ABL1* ratios were measured regularly during TKI reduction and after TKI cessation. Patients' remission status was reported for a 24 months follow-up or until disease recurrence. For most patients, additional data were available on *BCR-ABL1* measurements before trial entry.

*BCR-ABL1/ABL1* ratios were measured using reverse transcriptase quantitative (RTqPCR) techniques. At least 10,000 copies of control *ABL* transcripts were required for a valid result.

If no copies of *BCR-ABL1* are detectable in a blood sample, the measurement is indicated as non-detectable (ND). For the analysis in this paper, we calculate a detection limit for these measurements, applying the following formula:

$$\left(\frac{BCR\text{-}ABL1}{ABL1}\right)_{ND} = \frac{3}{ABL1 \cdot 100}$$

We use this value as an estimate of the detection threshold reasoning that the 'true' *BCR-ABL1/ABL1*-Ratio is lower.

Recurrence was molecularly defined as the time of the first of two consecutive measurements above 0.1% (i.e. loss of MR3).

The present study is based on 72 DESTINY entrants in whom additional data were available on *BCR-ABL1* measurements before trial entry. The complete time course data of those selected patients are composed of the full treatment response after diagnosis and TKI start, the 12 month reduction period and the follow-up period after stopping TKI (denoted as the *clinical reference data set*). To assess whether the model is able to make predictions by using incomplete data, we use also a *reduced data set* containing only the initial treatment response and the 12 month reduction period, while the follow-up period after TKI stopping has been truncated (see Supplementary Text S5).

#### S2: Mathematical modeling

In our established mathematical model of TKI-treated CML we explicitly consider immune effector cells (*Z*), quiescent (*X*) and proliferating (*Y*) leukemic stem cells (compare Figure 1B). The ordinary differential equation model is given by:

$$\frac{dX}{dt} = p_{YX} \cdot Y - p_{XY} \cdot X$$

$$\frac{dY}{dt} = p_{XY} \cdot X - p_{YX} \cdot Y + p_Y \cdot \left(1 - \frac{Y}{K_Y}\right) \cdot Y - m \cdot Z \cdot Y - d \cdot TKI \cdot Y$$

$$\frac{dZ}{dt} = r_z + Z \cdot p_z \cdot \frac{Y}{K_Z^2 + Y^2} - a \cdot Z$$

The model specifically distinguishes two functionally different states of leukemic stem cells (LSCs), namely a quiescent ($X$) and an actively cycling state ($Y$). Cells can reversibly switch between these states with transition rates $p_{XY}$ and $p_{YX}$. We assume that quiescent LSCs ($X$) do not proliferate, are insensitive to TKI effects, and do not interact with immune cells. In contrast, actively cycling LSCs ($Y$) proliferate with proliferation rate $p_Y$, while their proliferation is limited by a carrying capacity $K_Y$. Unlike quiescent LSCs, actively cycling cells are susceptible to TKI (described by the kill rate $TKI$) and immune effector cells (described by the kill rate $m$ per immune cell).

The immune compartment comprises a diverse pool of immune cells and other relevant factors responsible for the immunological interactions with leukemic cells, which potentially establishes a long-term control on treatment cessation. Immune cells ($Z$) are generated at a constant, low production rate $r_Z$ and undergo apoptosis with rate $a$. The leukemia-dependent recruitment of immune cells follows a nonlinear functional response where $p_Z$ and $K_Z$ are positive constants. This functional response leads to optimal immune cell recruitment, whereby at low proliferating leukemic cell levels ($Y < K_Z$), immune cell recruitment increases and immune cells $Z$ are stimulated to replicate in presence of proliferating leukemic cells $Y$, reaching a maximum $p_Z / (2\ K_Z)$ when $Y = K_Z$. For higher leukemic cell numbers $(Y > K_Z)$ the immune cell recruitment decreases with $Y$, reflecting the assumption that the proliferation of immune cells is decreased for high levels of proliferating leukemic cells $Y$ [2-4].

Assuming a linear dose-response relationship for the TKI treatment [5], the dosage factor *d* declines from $d=1$ (full dose) to $d=0.5$ or $0.25$ (reduced dose) to $d=0$ (therapy cessation).

**S3: Choice of parameters**

Model parameters

Within our model, a set of 11 parameters determines the resulting *BCR-ABL1/ABL1* time courses. As in a similar approach reported earlier [6], we chose fixed, universal values (i.e. applied to all patients in the same manner) for the immune mediated killing rate $m$, the carrying capacity $K_Y$, the immune cells natural influx $r_Z$ and the immune cells apoptosis rate $a$ that apply to all patients (see below). Furthermore, the following parameters are considered to be patient-specific: the initial *BCR-ABL1/ABL1*-ratio on the log-scale ($initLRATIO$), the transition rates $p_{XY}$ and $p_{YX}$, the TKI kill rate $TKI$, the proliferation rate $p_Y$ and the immune parameters $K_Z$ and $p_Z$.

*Parameter with fixed (universal) values*

For the fixed parameters, namely the immune mediated killing rate $m$, the carrying capacity $K_Y$, the immune cells natural influx $r_Z$ and the immune cells apoptosis rate $a$ we assign the following values: Given that an adult has about $10^6$ actively contributing stem cells [7], we define the combined carrying capacity $K_Y$ = $10^6$ of the Y-compartment regardless whether these cells are healthy HSC or LSC. The immune cells natural influx $r_Z$ and the immune cells apoptosis rate $a$ are chosen at $r_Z = 200$ *cells/month* and $a = 2$ $month^{-1}$, respectively, ensuring a constant low production level of immune cells. In order to ascertain a minimal basic immune activity, we use an immune mediated killing rate $m = 1*10^{-4}/cells*month$.

*Individually fitted parameters*

For each patient we apply a genetic algorithm to obtain optimal values for the initial *BCR-ABL1/ABL1*-ratio on the log-scale (*initLRATIO)*, the transition rates $p_{XY}$ and $p_{YX}$, the TKI kill rate *TKI*, the proliferation rate $p_Y$ and the immune parameters $K_Z$ and $p_Z$, to which we commonly refer to as the parameter set $\lambda = (initLRATIO, p_{XY}, p_{YX}, TKI, p_Y, K_Z, p_Z)$. For the optimization procedure we defined reasonable ranges for each parameter, which are given in Table ST 1 below.

Table ST1: Parameter ranges and parameter values. # *the value for* $p_Z$ *depends on both* $K_Z$ *and a. It is calculated as* $[0.5*a*2*10\text{^}K_Z^{Min}; N*a*2*10\text{^}K_Z^{Max}]$*, where N = 1000*

| Parameter | Ranges / values | Unit |
|---|---|---|
| $\boldsymbol{initLRATIO}$ | $10^1$ *to* $10^2$ | % |
| $\boldsymbol{TKI}$ | 0.5 *to* 3 | $month^{-1}$ |
| $\boldsymbol{p_{XY}}$ | $10^{-10}$ *to* 1 | $month^{-1}$ |
| $\boldsymbol{p_{YX}}$ | $10^{-10}$ *to* 1 | $month^{-1}$ |
| $\boldsymbol{p_Y}$ | 0.05 *to*$10^1$ | $month^{-1}$ |
| $\boldsymbol{K_Z}$ | $10^1$ *to* $10^{3.5}$ | cells |
| $\boldsymbol{p_Z}$ # | 20 *to* 12649111 | $cells^{-1}$ * $month^{-1}$ |
| $\boldsymbol{K_Y}$ | $10^6$ | cells |
| $\boldsymbol{m}$ | $10^{-4}$ | $cells^{-1}$ * $month^{-1}$ |
| $\boldsymbol{rz}$ | 200 | cells*$month^{-1}$ |
| $\boldsymbol{a}$ | 2 | $month^{-1}$ |

Genetic algorithm

For the identification of optimal model parameters for each patient we use a *genetic algorithm* (R version 3.6.3, package rgenoud version 5.8-3.0). In brief, a genetic algorithm mimics a search strategy inspired by evolutionary biology, in which the fitness of a particular parameter choice (in terms of an objective function, which in our case is the residual error between model and data) determines its chance of 'survival'. While the fittest 'individuals' reproduce, their parameterization is altered and reproduced among a larger number of progeny, for which the same rules for 'survival' apply. Alteration of the parameters are initiated via different operators, like *mutation*, *crossing over* or *cloning* which follow concepts of evolutionary biology. This

process is repeated for multiple generations until the results do not ameliorate anymore. Unlike many classical optimization routines, genetic algorithms are suited to broadly scan a large search space in which many local minima exist.
Multiple initializations of the optimization routine can lead to slightly different results, indicating that sparse and intrinsically noisy measurements limit the identifiability of a uniquely optimal parameter set. Instead of focusing on one ("most") optimal parameter set, we initiate the optimization routine multiple times (in our case $i \in \{1 \dots 64\}$) and take the resulting set of 64 optimal parameterizations $\lambda_{ij} = \left(initLRATIO, p_{XY}, p_{YX}, TKI, p_Y, K_Z, p_Z\right)$, as a surrogate to approximate the range of plausible models for each patient *j*.

### S4: Sampling approach to estimate the timing and fraction of recurrences

In order to compare different treatment scenarios, we estimate the fraction of expected recurrences as a function of time (cumulative incidence). As we describe each patient by a set of 64 optimal parameterizations $\lambda_{ij}$, we derive the corresponding estimations by the following sampling strategy: For each patient *j* we randomly choose one eligible parameter set $\lambda_{ij}$. Doing so for each patient we obtain a sample of 72 single parameter sets mimicking the complete patient cohort. Using these parameter sets, we simulate the outcome for a particular treatment scheme on a per patient basis from which a cumulative incidence curve is derived. Repeating this sampling process 1000 times, we obtain a spectrum of potential realizations that reflects the inherent heterogeneity which is also contained in the non-unique parameter estimates $\lambda_{ij}$ for each patient. We determine the median and a 95%-confidence region over this spectrum of realizations to obtain the cumulative incidence curves shown in Figures 2, 4 and 5.
To further examine how treatment duration acts on the different immune classes (compare Supplementary Text S6), we stratified the patients as to whether any of their optimal parameter sets $\lambda_{ij}$ belong to class A, B or C. The approach is now restricted to the patients with at least one fit of the corresponding immune class, while the sampling only considers that fits corresponding to the respective immune class. At each repetition, one of the available fits is randomly chosen to represent this patient in the sampling procedure.

### S5: Recurrence prediction for reduced time course data

We explored whether our modeling approach can also be trained on pre-cessation data only while maintaining its correct description of the remission behavior after therapy stop. To this end, we apply the model to a reduced data set containing only the initial treatment response and the 12 month reduction period, while the follow-up period after TKI stopping has been truncated (*reduced data set*, see Supplementary Text S1). We then compare the simulated future time courses to the reference simulation which was optimized on the full *clinical reference data set.*

Technically, we apply the same optimization routine to obtain another 64 parameter configurations $\lambda^*_{ij}$ for each patient *j* with the *reduced data set* only (indicated by * notation). Visual comparison of the simulation to the clinical data (see example in Figure S4A) confirms that the dynamics of initial treatment response and during dose reduction are well captured. In a next step, time courses are simulated for each parameter set $\lambda^*_{ij}$, beyond the time point of treatment cessation. Applying the same binary classification for recurrence detection (*simulated recurrence patient* vs. *simulated TFR patient*), we compared the extended model simulations with the clinical outcome. The summary in Figure S4B, as well as the particular example shown in Figure S4A, indicate that the reduced data set is not sufficient to optimally match the clinical findings: only 29 out of 35 clinical TFR cases are correctly identified (indicating 6 false positives), while 22 out of 37 clinical recurrences are correctly predicted (indicating 15 false negatives). Figure S4C confirms that the simulated recurrences appear early after TKI stop, while the fraction of recurrences is also seriously underestimated using this complementary approach.

We previously identified that almost all DESTINY patients with a fast increase in *BCR-ABL1* levels during dose reduction (linear slope > 0.068 log[BCR-ABL1/ABL1] increase per month) have molecular recurrence after TKI cessation[8]. However, among the patients with non- or slowly-increasing *BCR-ABL1* levels during the dose reduction period (slope < 0.068 log[BCR-ABL1/ABL1] increase per month) 27% developed molecular recurrence by 24 months after therapy cessation [8]. Applying the same binary classification to the data set of 72 patients, we confirm those earlier findings in that the model correctly predicts recurrences for all patients with fast *BCR-ABL1* increase (high slopes in Figure S4D) during the dose reduction period, while this is not the case for patients with non- or slowly-increasing *BCR-ABL1* levels (low slopes in Figure S4D).

We speculate that *BCR-ABL1* levels that rise fast during TKI dose reduction might be identifying insufficient immune control in good molecular remission. However, predictions for patients with non- or slowly-increasing *BCR-ABL1* levels during dose reduction are inherently more difficult, as an inference of functional immunological control cannot be obtained with sufficient precision. It is particularly this subcohort of patients who require a better understanding of the mechanisms leading to loss of TFR.

### S6: Immune classes and calculation of steady states

The three immune classes A, B and C, to which a parameter set $\lambda_{ij}$ can be classified, are determined by the number and stability of steady states within their immune landscape (compare Figure 3A). Immune classes are calculated for the treatment free situation, i.e. $d = 0$. As we have not obtained a uniquely identifiable parameter set for all patients $j$, but use a spectrum of $64$ optimization runs $i$, optimal parameter sets $\lambda_{ij}$ of the same patient may be classified to different immunological classes A, B or C.

Parameter sets belonging to *class A* are characterized by generating an insufficient immune reaction to CML. In this case, as long as one single CML cell survives, leukemic cells will regrow in an uncontrolled manner and reach the only stable steady state describing leukemic dominance ($E_{High}$). Parameter sets belonging to *class B* refer to scenarios with a strong immune system, allowing for the coexistence of two steady states: while one steady state describes leukemic dominance ($E_{High}$, similar to class A patients), the second steady state is described by a sufficiently low number of leukemic cells, which are controlled by the activated immune effectors cells (remission steady state, $E_{Low}$). The third class of parameter sets, *class C*, presents with a similar immunological landscape as class B, with the same steady states. However, due to the size and the shape of the basin of attraction of steady state $E_{Low}$ (see

Figure 1C), the equilibrium between immune effector cells and leukemic cells is more difficult to achieve and requires a balance between stimulation of immune cells by leukemic cells and elimination of leukemic cells by the respective immune cells.

Following a more formal approach we point out that there exists a further steady state characterized by the absence of LSC, to which we refer as the complete eradication steady state ($E_0$). This steady state is distinct from the immunologically controlled remission steady state ($E_{Low}$), which has low LSC counts and high (enough) immune cell levels controlling the residual disease.

**(1) Eradication steady state ($E_0$)**

The complete eradication steady state is the trivial equilibrium $E_0 = (X_0, Y_0, Z_0)$ of the ODE-system. It is defined by

$$X_0 = 0$$

$$Y_0 = 0$$

$$Z_0 = \frac{r_Z}{a}$$

**(2) Immunological controlled remission and recurrence steady states**

In contrast to the eradication steady state, both the remission and the recurrence steady states are characterized by existent LSC counts. The immunological controlled remission steady state $E_{Low} = (X_L, Y_L, Z_L)$ presents with low levels of LSC while the recurrence steady state $E_{High} = (X_H, Y_H, Z_H)$ is indicated by high LSC counts i.e., $X_L < X_H$ and $Y_L < Y_H$.

The steady states are obtained as follows. Setting $dX/dt = 0$ and isolating $X$ we obtain

$$X = \left(\frac{p_{YX}}{p_{XY}}\right) \cdot Y$$

Analogously, setting $dZ/dt = 0$ and isolating $Z$ we obtain

$$Z = \frac{r_Z \cdot \left({K_Z}^2 + Y^2\right)}{a \cdot \left({K_Z}^2 + Y^2\right) - p_Z \cdot Y}$$

Now, setting $dY/dt = 0$ and substituting the expressions of $X$ and $Z$ we obtain, after simplification, the following third-degree polynomial equation in $Y$:

$$-a\,p_Y Y^3 + \left(p_Y p_Z + K_Y(a\,p_Y - m\,r_z)\right)Y^2 + -p_Y\left(aK_Z{}^2 + K_Y p_Z\right)Y + K_Y K_Z{}^2(a\,p_Y - m\,r_Z) = 0$$

Using the same techniques as in our previous publication [6] one can show that this equation does not have a positive solution $Y$ for $a \cdot p_Y < m \cdot r_Z$. In this case, both the remission $E_{Low}$ and recurrence $E_{High}$ steady states do not exist. On the contrary, there are two options for $a \cdot p_Y > m \cdot r_Z$, depending on the discriminant of equation above: either the equation has one or three positive solutions for $Y$. We omit the formula of the discriminant for sake of brevity. If the equation has three positive solutions, we denote them as $Y_H > Y_S > Y_L$. Substituting these values in the expressions of $X$ and $Z$ above, we obtain three equilibrium points: the recurrence steady state $E_{High}$, an intermediated steady state $E_S$, which is a saddle-point and separates the basins of attraction of $E_{High}$ and $E_{Low}$, and the remission steady state $E_{Low}$. For all parameter values used here, $E_{High}$ and $E_{Low}$ are stable fixed points, while $E_S$ and $E_0$ are saddle-points. Such configuration corresponds either to class B or class C. The distinction of such classes is made by assessing whether a system solution starting as small perturbation of $E_0$ will converge either to $E_{Low}$ or to $E_{High}$. Finally, if the above polynomial equation has only one positive solution $Y > 0$, then it defines solely the recurrence steady state $E_{High}$ while the remission steady state does not exist. In this case, we verify that $E_{High}$ is stable. This third scenario corresponds to class A.

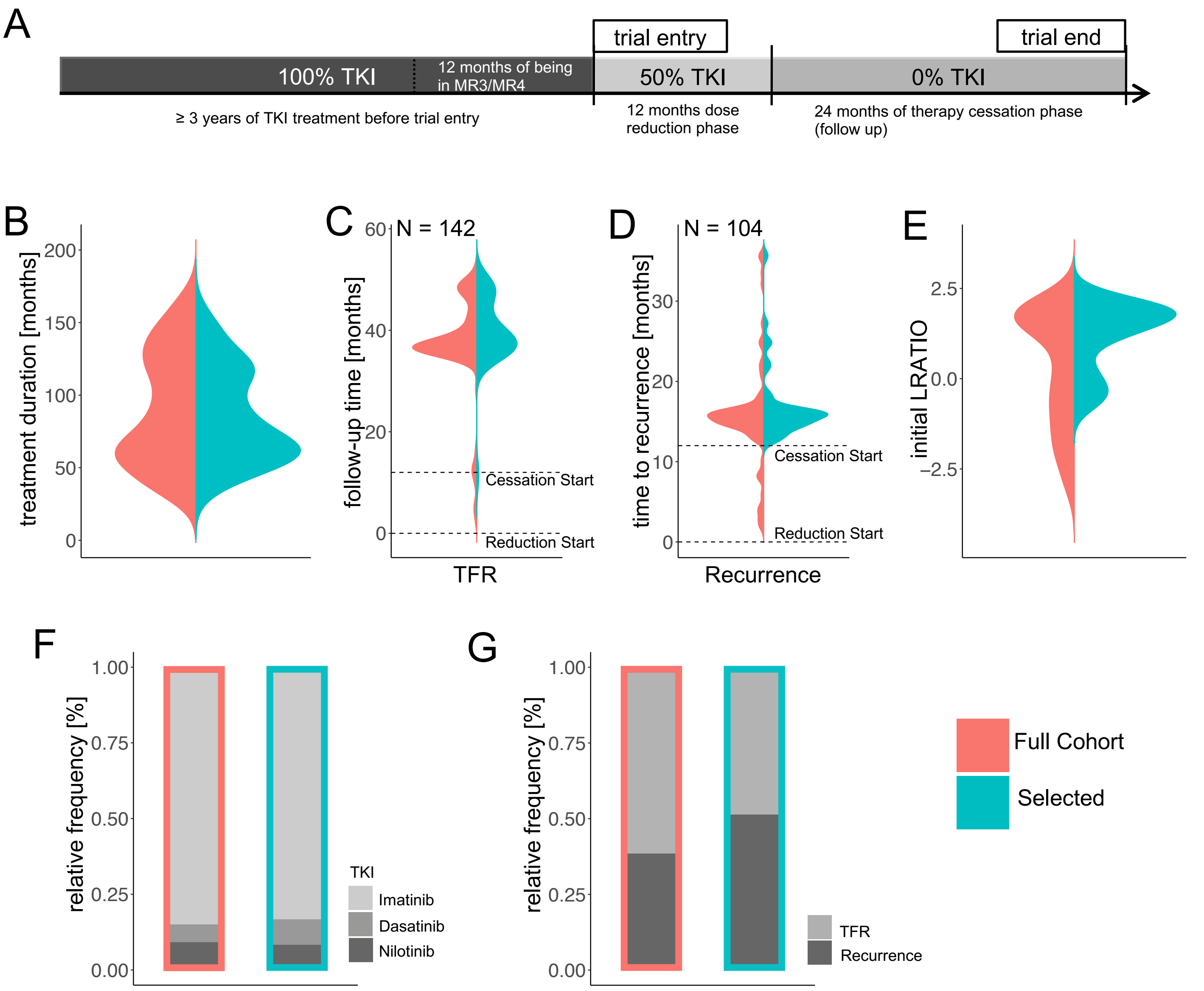


**Supplementary Figure S1:**

**A:** Time schedule of the DESTINY trial.

Comparison between original DESTINY cohort (N = 174, red) and the selected cohort (N = 72, green) with respect to:

**B**: TKI treatment duration until inclusion into DESTINY, i.e. start of dose reduction (For patients in the full cohort, for which the previous TKI treatment duration could not be unambiguously verified, this value was set 36 months.)

**C**: Follow-up time, indicating the end of follow up, measured from the start of dose reduction.

**D**: Time to recurrence, indicating the time point of recurrence, measured from the start of dose reduction.

**E:** Initial BCR-ABL1 ratio (on log scale, LRATIO=$\log_{10}$(BCR-ABL1/ABL1)).

**F**: Relative frequencies of used TKI.

**G**: Relative frequencies of recurrences.

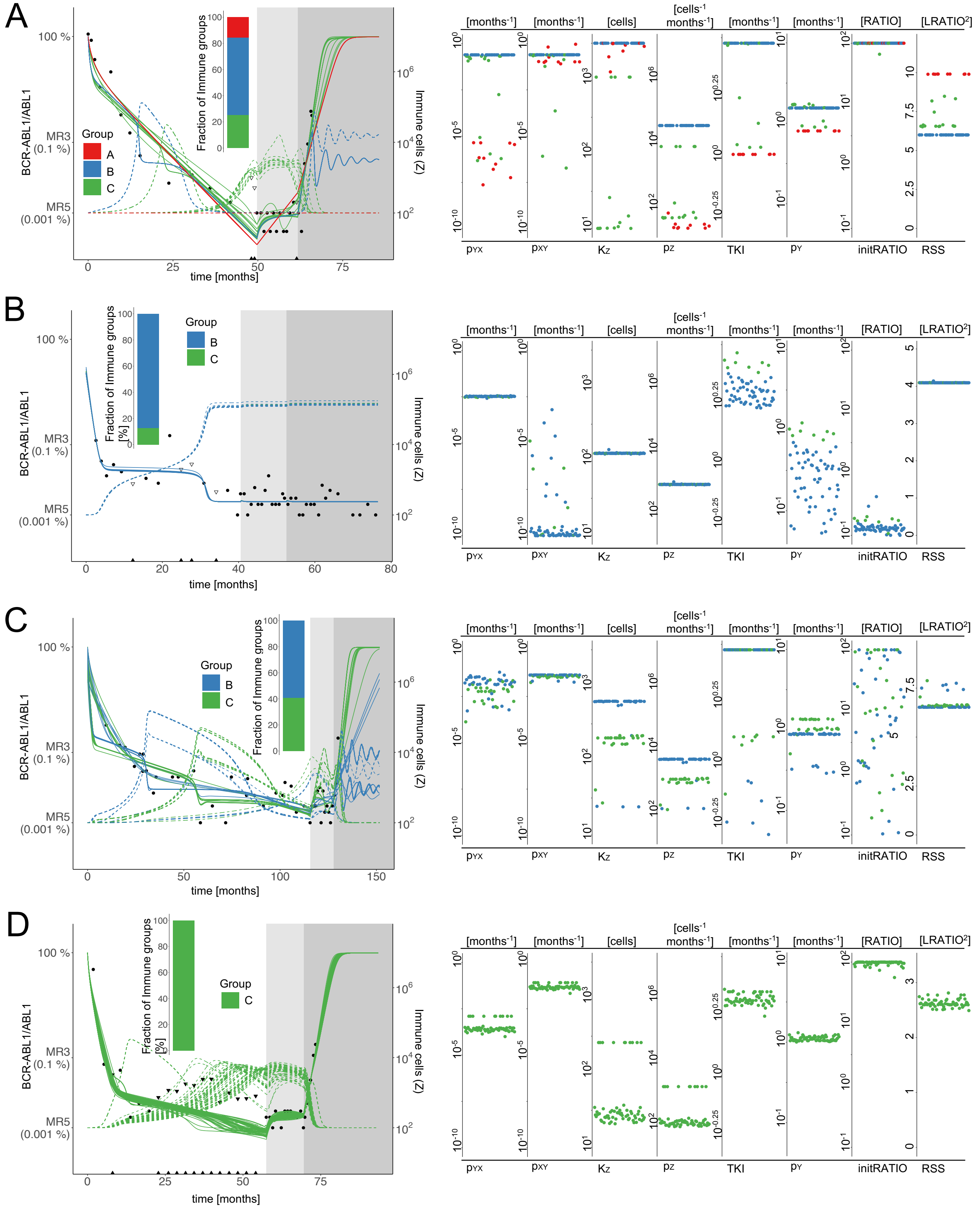


**Supplementary Figure S2: A-D**

**(left)** Example of clinical data and model simulations of 4 patients, patients A & C with CML recurrence, patients B & D with TFR after therapy stop. Black dots indicate BCR-ABL1/ABL1-measurements; black triangles indicate the quantification limit for undetectable BCR-ABL1 levels. White background indicates full dose TKI treatment, light grey indicates the reduction period (50% of full dose), and dark grey refers to the time period after treatment cessation. 64 simulated BCR-ABL1/ABL1-time courses (solid lines) and corresponding simulated immune cell counts (dashed lines) are colored according to their immune classification (red – A, blue - B, green - C). The inset shows the distribution of the immune classes within this patient.

**(right)** Distribution of individual parameter values of the 64 parameter sets of the same patients as on the left side, shown separately for the 7 free parameters along with the residual sum of squares (RSS) as a measure of fitting quality. Fits are colored according to the immune classification of the corresponding parameter set.

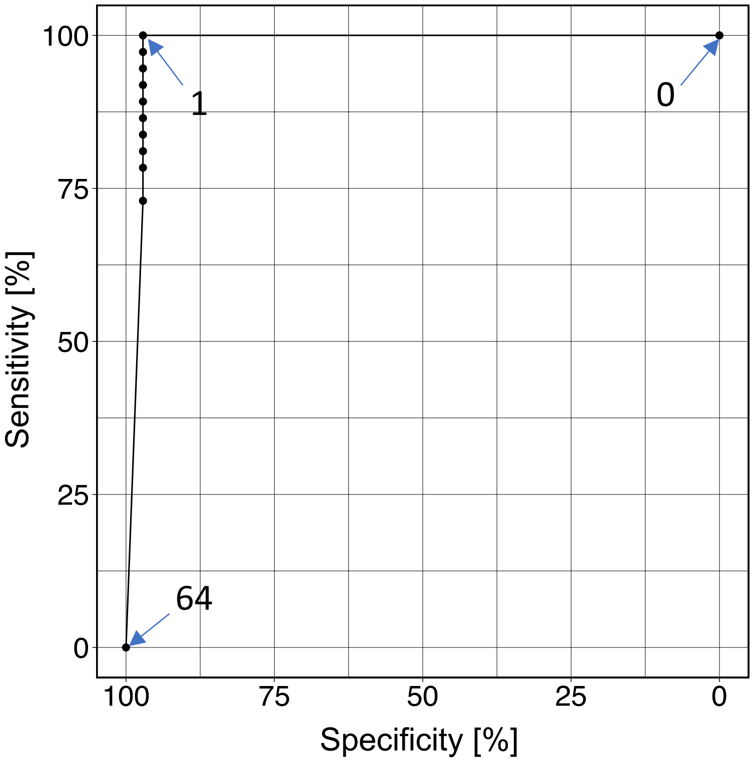


**Supplementary Figure S3:**
Sensitivity as a function of the false-positive-rate for different thresholds for the classification of patients to either the TFR-group or the recurrence-group (ROC-Curve). Dots indicate the different thresholds. A threshold of x means: if x or more parameter sets $\lambda_{ij}$ indicate a recurrence, the whole patient is classified into the recurrence group.

A

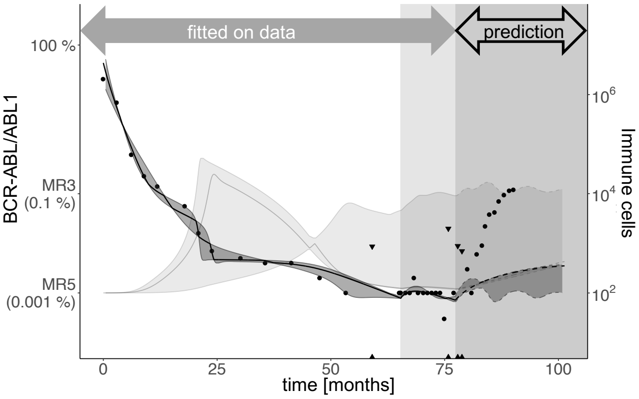


B

All patients

| N = 72 | clinical recurrence | clinical TFR |
|---|---|---|
| **simulated recurrence** | **22** (30.6%) | **6** (8.3%) |
| **simulated TFR** | **15** (29.8%) | **29** (40.3%) |

C

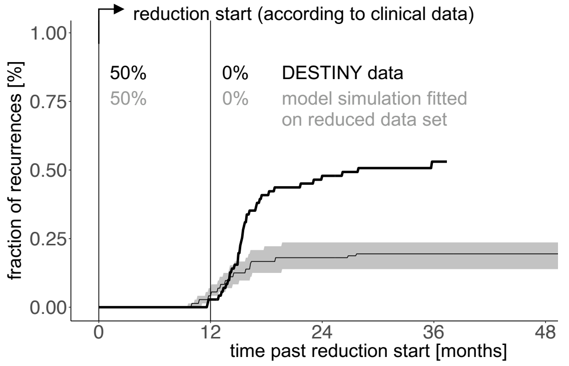


D

High slope

| N = 15 | clinical recurrence | clinical TFR |
|---|---|---|
| **simulated recurrence** | **14** (93.3%) | **1** (6.7%) |
| **simulated TFR** | **0** | **0** |

Low slope

| N = 57 | clinical recurrence | clinical TFR |
|---|---|---|
| **simulated recurrence** | **8** (14.0%) | **5** (8.8%) |
| **simulated TFR** | **15** (26.3%) | **29** (50.9%) |

**Supplementary Figure S4**:
**A)** The simulation model is only fitted on the clinical data before treatment cessation (reduced data set: the fitting procedure only considers data during full dose treatment (white background) and dose reduction (light grey background). The dashed lines after treatment cessation (dark grey background) indicate predictions based on the model simulations along with the corresponding confidence regions. While the clinical data indicates a disease recurrence, the model does not reflect this behavior for the selected example.
**B)** Comparison of the simulated results (i.e. binary classification into *simulated recurrence* and *simulated TFR*) and the clinical outcomes for the case that the fitting is only applied to clinical data before treatment cessation.
**C)** Comparison of the time and fraction of recurrences between the clinical data (black line) and the model prediction based on fitting to the reduced data set (grey). To obtain the curve for the simulation scenario, we randomly choose one eligible parameter set $\lambda_{ij}$ per each patient *j* and calculate whether and when there will be a recurrence in the corresponding simulation. We repeat this sampling approach 1000 times to obtain the median (grey line) and a 95%-confidence region (grey shaded).
**D)** The comparison in B) is shown separately for patients with strongly increasing *BCR-ABL1* levels during dose reduction (linear slope > 0.068 log[BCR-ABL1/ABL1] increase per month (High slope), N=15) and non- or slowly-increasing levels (slope < 0.068 log[BCR-ABL1/ABL1] increase per month (Low slope), N=57).

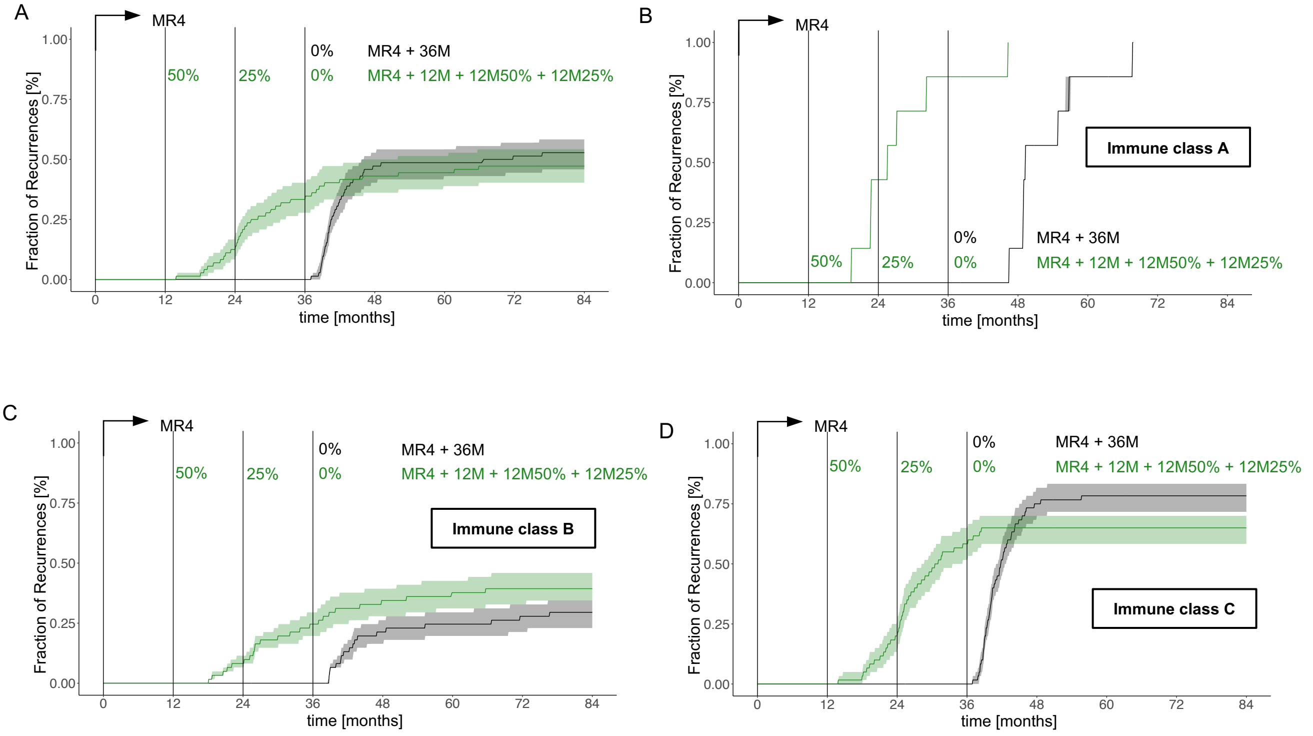


**Supplementary Figure S5:**
Fraction of recurrences as a function of time comparing immediate therapy cessation 36 months after reaching MR4 (black) with a treatment scenario with 12 months full dose treatment past reaching MR4 plus 12 month dose reduction to 50% of the initial dose plus 12 month dose reduction to 25% of the initial dose (green, indicated as MR4 + 12M + 12M 50% + 12M 25%)
The 4 plots compare different subcohorts of the analysed fits:
**A)** Combined results over all fits (see Fig. 5D)
**B)** Sampling over all fits that belong to immune class A
**C)** Sampling over all fits that belong to immune class B
**D)** Sampling over all fits that belong to immune class C

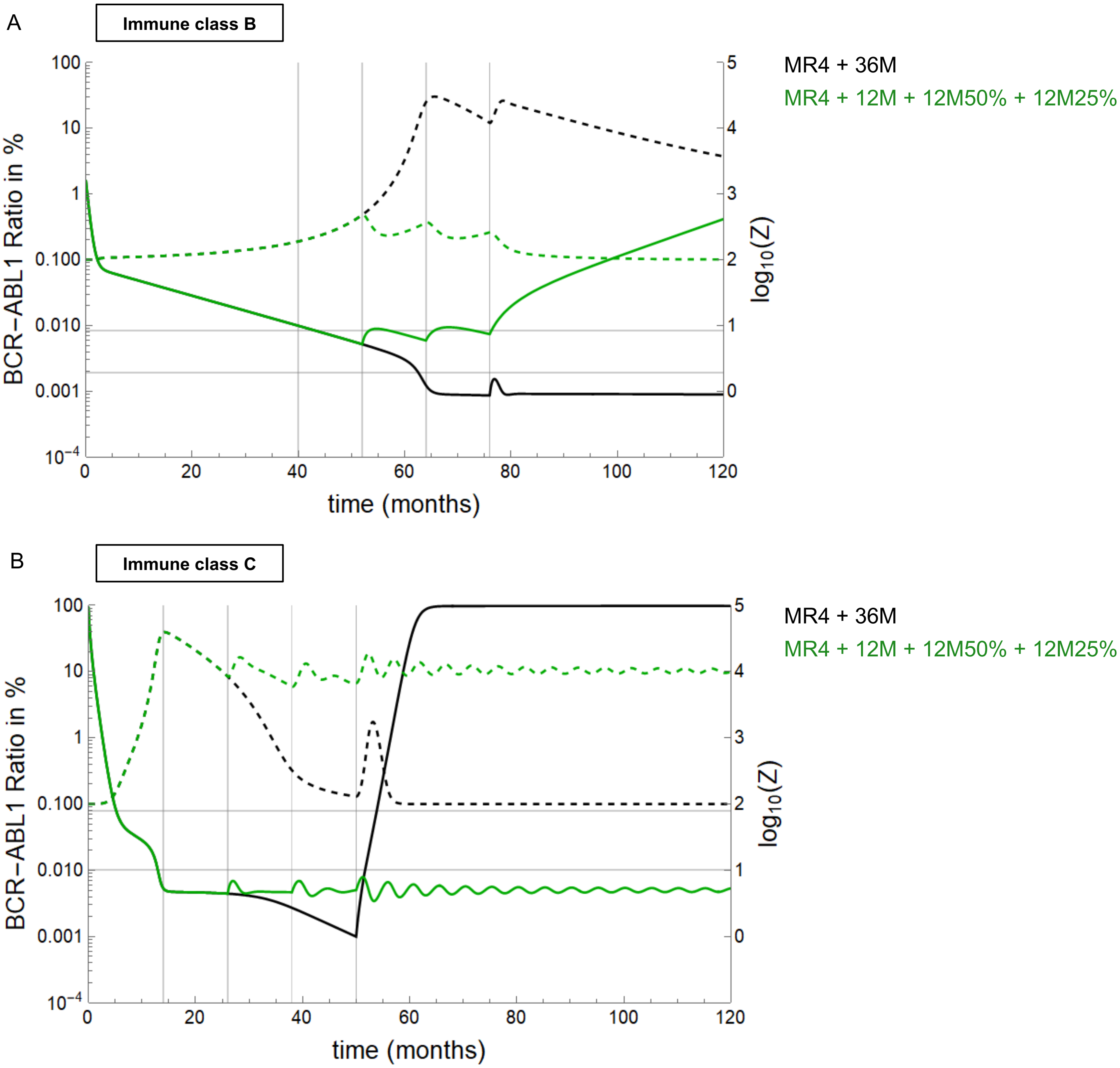


**Supplementary Figure S6:**
Model simulations of two treatment schemes (MR4+12M+12M50%+12M25%, green; and MR+36M, black) for two representative patients belonging to class B (**panel A**) and class C (**panel B**). Continuous lines represent *BCR-ABL1 ratios*, while dashed lines represent immune cell numbers (Z). These two individual simulations illustrate the outcome observed on the population level (Supplementary Figure S5 C,D): while the class C patient benefits from a stepwise reduction scheme (green) and achieves TFR, the opposite outcome occurs for the class B patient that does not reach the remission state.

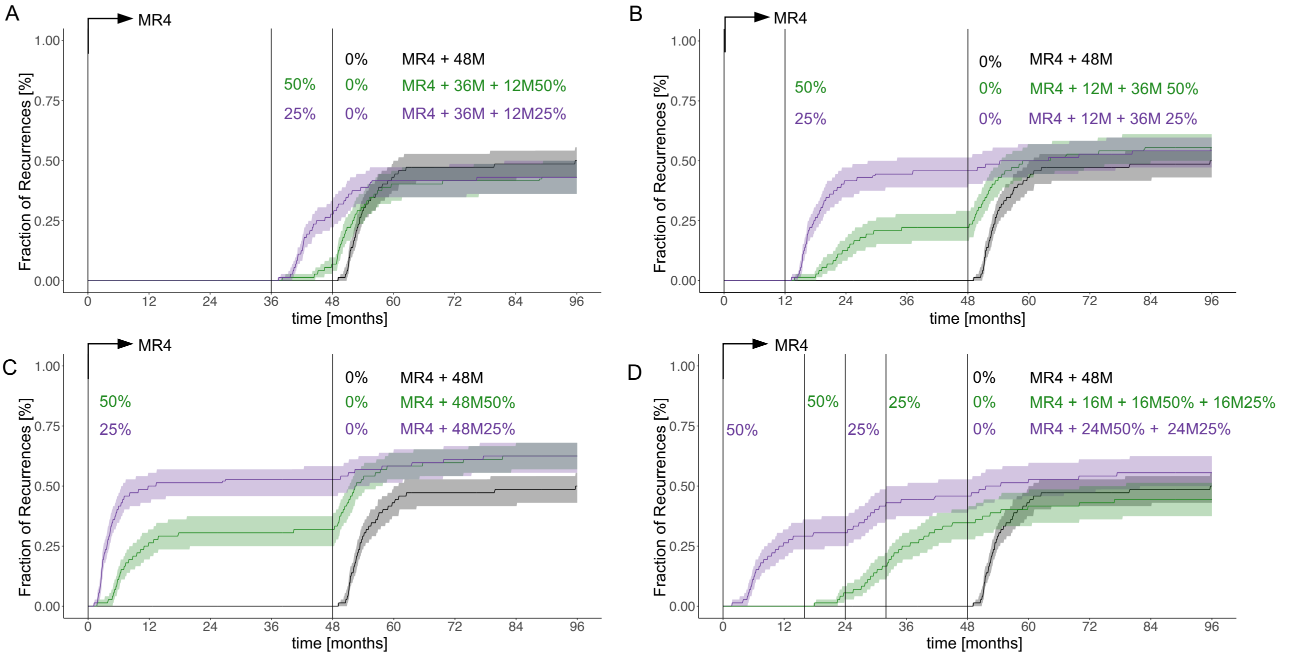


**Supplementary Figure S7:**
Fraction of recurrences as a function of time comparing immediate therapy cessation 48 months after reaching MR4 (black) with different scenarios of dose reduction covering the same overall treatment duration:
**A)** 36 months full dose treatment past reaching MR4 plus 12 month dose reduction to 50% of the initial dose (green, indicated as MR4 + 36M + 12M 50%) and 36 months full dose treatment past reaching MR4 plus 12 month dose reduction to 25% of the initial dose (purple, indicated as MR4 + 36M + 12M 25%).
**B)** 12 months full dose treatment past reaching MR4 plus 36 month dose reduction to 50% of the initial dose (green, indicated as MR4 + 12M + 36M 50%) and 12 months full dose treatment past reaching MR4 plus 36 month dose reduction to 25% of the initial dose (purple, indicated as MR4 + 12M + 36M 25%).
**C)** dose reduction to 50% of the initial dose for 48 months immediately after reaching MR4 (green, indicated as MR4 + 48M 50%) and dose reduction to 25% of the initial dose for 48 months immediately after reaching MR4 (purple, indicated as MR4 + 48M 25%).
**D)** 16 months full dose treatment past reaching MR4 plus 16 month dose reduction to 50% of the initial dose plus 16 month dose reduction to 25% of the initial dose (green, indicated as MR4 + 16M + 16M 50% + 16M 25%) and dose reduction to 50% of the initial dose for 24 months immediately after reaching MR4 plus 24 month dose reduction to 25% of the initial dose (purple, indicated as MR4 + 24M 50% + 24M 25%).
